# Internal medicine clerkship amidst COVID-19 pandemic: clinical learning experience of undergraduate medical students at Makerere University, Uganda: a cross-sectional study

**DOI:** 10.1101/2020.12.16.20248110

**Authors:** Felix Bongomin, Ronald Olum, Lydia Nakiyingi, Lalitha Rejani, Isaac Ssinabulya, Christine Sekaggya-Wiltshire, Ponsiano Ocama, Pauline Byakika-Kibwika

**Affiliations:** Department of Medicine, College of Health Sciences, Makerere University, Kampala, Uganda; Department of Medical Microbiology and Immunology, Faculty of Medicine, Gulu University, Gulu, Uganda; Uganda Heart Institute, Mulago National Referral Hospital, Kampala, Uganda; Directorate of Medicine, Mulago National Referral Hospital, Kampala, Uganda

**Keywords:** Clinical Learning Experience, COVID-19, Internal Medicine, Makerere University

## Abstract

**Background:** The coronavirus-2019 (COVID-19) pandemic continues to impose a significant impact on medical education. We aimed to describe the clinical learning experience of undergraduate medical students undertaking internal medicine clerkship during the COVID-19 pandemic at Makerere University, Uganda.

**Methods:** A descriptive, cross-sectional study among medical students in clinical years of study pursuing the Bachelor of Medicine and Bachelor of Surgery undergraduate degree program was conducted in November 2020. Only 3^rd^ (junior clerks) and 5^th^ (senior clerks) year medical students whose internal medicine clerkships were interrupted by the COVID-19 pandemic were studied.

**Results:** Data of 188 (95%) eligible clinical year students; junior (101, 54.0%) and senior (86, 46.0%) were analysed. Median age was 24 (range: 22-42) years. Majority (70.1%) were male and Ugandan nationals (94.1%). Sixty-four (30.3%) students reported inadequate personal protective equipment, 152 (81.7%) felt at risk of contracting COVID-19, and 127 (67.9%) said it was difficult to observe COVID-19 standard operating procedures. Twenty-two students (11.9%) were discouraged from pursuing a career in internal medicine. Overall, most students reported good or excellent clinical experience pre-COVID-19 era compared to during the COVID-19 era (4.0 vs. 3.5, p<0.0001). Senior clerks significantly believed that the time allocated for the rotation was adequate (p<0.0001) and they were able to complete their study objectives (p<0.001), compared to the junior clerks. Senior clerks believed that learning was difficult when combined with junior clerks (p=0.013). About half of the students (51.4%, n=95) reported clinical teaching should remain as it was in the pre-COVID-19 era.

**Conclusions:** The COVID-19 pandemic has had a significantly negative effect on the clinical learning experience of the students. There is need to review the current teaching and learning methods to suit teaching and learning during pandemics of highly infectious diseases to ensure safe and effective learning experience.

**Highlights:**

1. Nearly half of the medical students reported a negative clinical learning experience during their internal medicine clerkship in the COVID-19 pandemic.
2. Over two-thirds of the participants expressed that it was difficult to observe COVID-19 prevention standard operating procedures during their clinical rotations.
3. Over 80% of the medical students felt at risk of contracting COVID-19 during their clinical rotation.
4. About one-third of the medical students reported inadequacy in personal protective equipment like masks during their rotation.

## INTRODUCTION

The on-going coronavirus disease -2019 (COVID-19) pandemic, caused by the novel severe acute respiratory syndrome-coronavirus-2 (SARS-CoV-2) continues to be a global crisis with devastating effects across all sectors, including healthcare and medical education [1,2]. Within a year, the highly infectious SARS-CoV-2 virus has rapidly spread in all continents of the world infecting over 74 million persons resulting in approximately 1.6 million deaths worldwide [3]. Preventive approaches such as physical distancing, wearing of face masks, and observing respiratory and hand hygiene remains the mainstay of averting the spread of the virus [4–6].

Healthcare systems across the world have been over-stretched by the increasing number of patients with COVID-19 [7]. In fact, in some countries hit-hard by the pandemic, medical students in their final clinical year of medical training were fast-tracked to graduate to boost the existing healthcare workforce [8]. Medical students are experiencing this unique training environment for the first time during their clinical training. The impact of this situation on their acquisition of clinical knowledge and skills and how this may affect their future career choices remains elusive.

In Uganda, as lockdown was being eased, following standard operating procedures (SOPs) outlined by the Ministry of Health, medical students in their clinical years of training at Makerere University were allowed back on the hospital wards to continue with their clinical learning in early October 2020 [9].

As the number of COVID-19 cases in Uganda continues to exponentially increase in excess of 28,168 cases with over 225 deaths including frontline healthcare workers [3], the fear of contracting the disease from patients may have a significant effect on the learning experience of medical students. The teachers were concerned that students’ acquisition of clinical skills competence may be compromised as patient access is limited and some of the routine clinical examination procedures could be affected by the standard operating procedures for preventing COVID-19 transmission.

The clinical learning experience of the medical students during the pandemic in Uganda is poorly understood and not documented. We therefore aimed to describe the clinical learning experience of medical students who undertook internal medicine clerkship during the pandemic at Makerere University, Uganda.

## MATERIALS AND METHODS

### Study design and period

This was a descriptive, cross-sectional study conducted in the last week of November 2020.

### Study settings

The study was conducted at the Department of Medicine, School of Medicine, Makerere University College of Health Sciences (MakCHS). MakCHS is located at Mulago Hospital Complex, Mulago Hill, 5 kilometres North-East of Kampala, Uganda’s Capital. MakCHS has four schools offering 12 undergraduate degree programs with a population of approximately 2,000 students as of Academic Year 2019/2020. The duration of study ranges from 3 years to 5 years depending on the courses offered. Students on the Bachelor of Medicine and Bachelor of Surgery (MBChB) degree program take a minimum of 5 years to complete their training. There are about 200 5^th^ year (senior clerks) and 180 3^rd^ year (junior clerks) students in the academic year 2019/2020 undertaking the MBChB program. The students for junior and senior clerkship rotations are divided into 4 groups, with each semester having 2 groups rotating in internal medicine each for a period of 7.5 weeks.

### Study population

All 3^rd^ and 5^th^ Year MBChB students who undertook the internal medicine rotation during the October –November 2020 clinical clerkship period were included in the study after informed consent was obtained.

### Data collection

We employed purposive sampling. For the quantitative study, paper-printed semi-structured questionnaires were self-administered. Data was collected immediately after candidates wrote their final internal medicine theory exams. In all settings during data collection, COVID-19 standard operating procedures (SOPs) as outlined by Ministry of Health, Uganda [9] were observed.

### Study variables

Independent variables were demographic characteristics including sex, age, year of study, and nationality. Dependent variables were teaching and learning experience and challenges to clinical clerkship during COVID-19 pandemic. A 5-point Likert scale (very poor [1], poor, fair, good and excellent [5] OR strongly agree [5], agree, neutral, disagree and strongly disagree [1]) was used to evaluate overall learning experience of the learners during the study period. The study questionnaire was adapted from a previously validated and published tool [10].

### Statistical analyses

Data from completed questionnaires were entered into Microsoft Excel 2016 for cleaning and coding and later exported to STATA version 16.0 for analyses. Numerical data were summarized as means and standard deviations if normally distributed and medians and ranges if non-normally distributed. Categorical data were summarized as frequencies and percentages. Associations between independent variables and dependent variables were assessed using parametric or non-parametric tests and multivariable analysis as appropriate. Internal reliability of each of the questions was assessed using Cronbach alpha reliability test. A *P*<.*05* was considered statistically significant.

## RESULTS

### Characteristics of participants

All 201 eligible medical students completed the survey (response rate= 100%). However, 13 participants had incomplete questionnaires and were therefore excluded from the final analysis.

Overall, data of 188 medical students with a median age of 24 (interquartile range: 21 to 40) years were analysed (**Table 1**). Majority of the participants were Ugandans (94.2%, n=177), male (70.7%, n=133) and residing in university halls (50%, n=94). About 72% (n=161) of the participants got food from nearby restaurants and the median daily expenditure on food was 5,000 Uganda Shillings (∼1.4 US dollars (USD)). Only 110 (58.5%) medical students used the bus provided by the university whereas the others spent a median amount of 10,000 UGX (∼2.3 USD) daily on transport to the hospital.

**Table 1.**
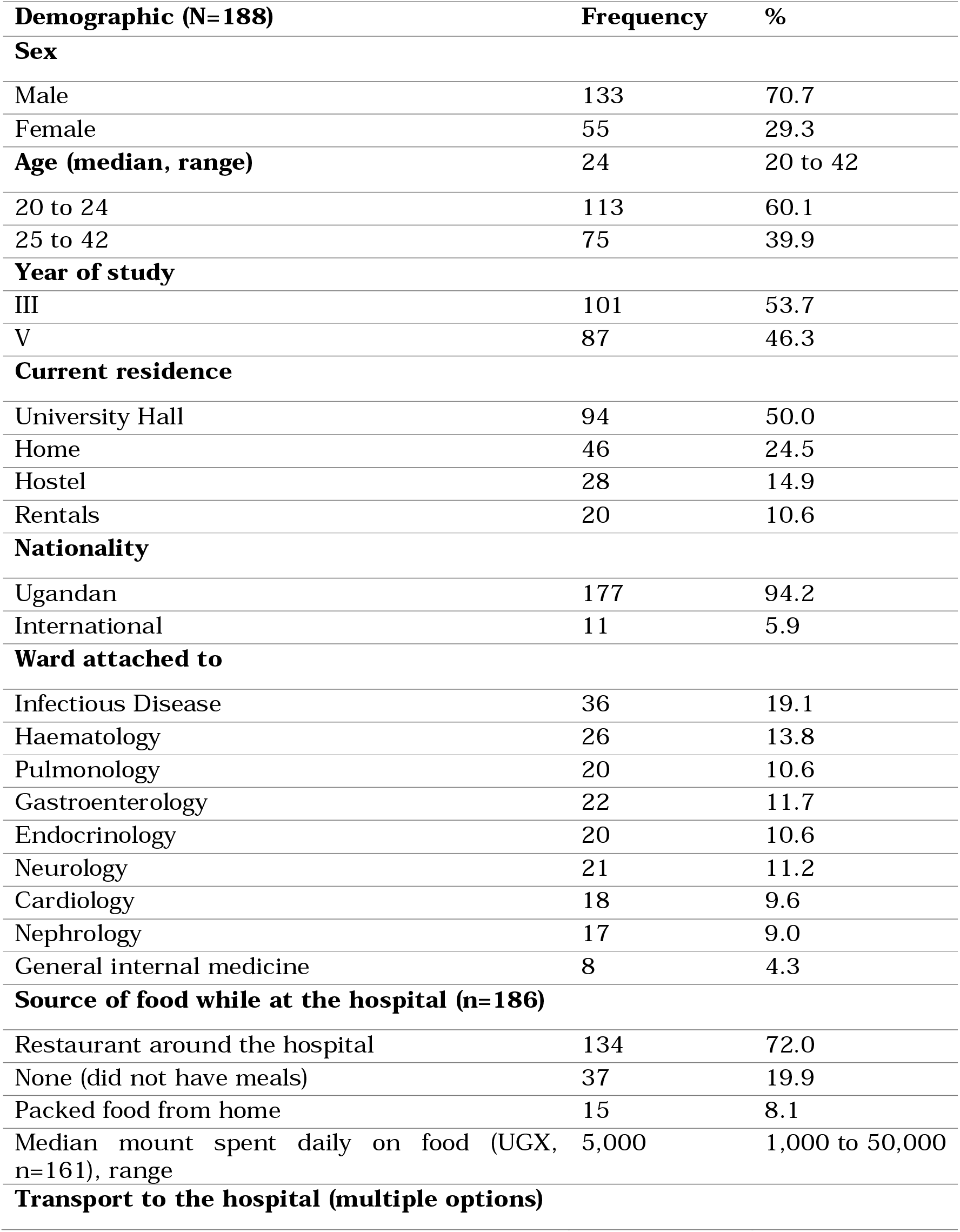
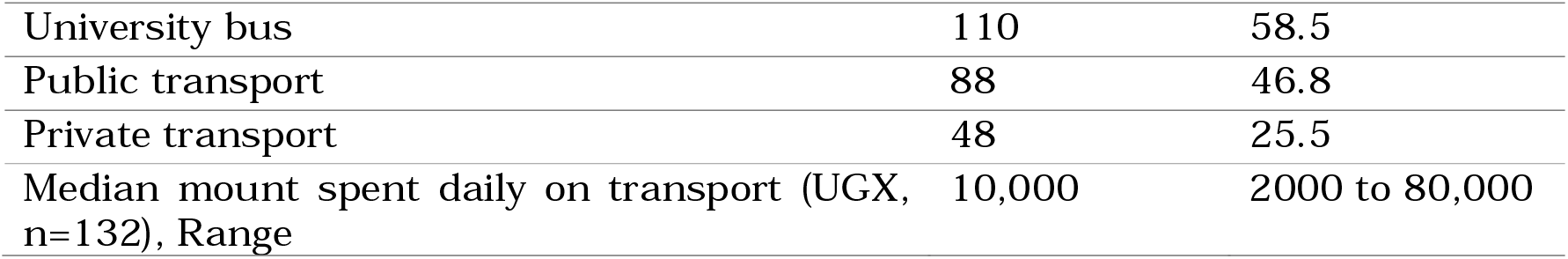
Social and demographic characteristics of the participants

### Learning experience during the COVID-19 pandemic

**Table 2** summarizes the experiences of medical students during internal medicine rotation amidst the pandemic. The mean experience scores for most items assessed were ≥4 indicating good experiences for the respective items assessed (**Table 3**).

**Table 2.** Learning experiences of medical student during internal medicine clerkship amidst COVID-19 pandemic.

| Experiences (Cronbach = 0.78) | n | Excellent,<br>n (%) | Good,<br>n (%) | Fair,<br>n (%) | Poor,<br>n (%) | Very Poor,<br>n (%) |
| --- | --- | --- | --- | --- | --- | --- |
| Clinical supervision by university lecturers | 184 | 43 (23.4) | 112 (60.9) | 23 (12.5) | 5 (2.7) | 1 (0.5) |
| Patient-centred learning | 186 | 76 (40.9) | 80 (43) | 27 (14.5) | 2 (1.1) | 1 (0.5) |
| Team work | 187 | 69 (36.9) | 99 (52.9) | 18 (9.6) | 0 (0) | 1 (0.5) |
| Learning efforts | 183 | 44 (24) | 112 (61.2) | 26 (14.2) | 1 (0.5) | 0 (0) |
| Quality of teaching | 184 | 41 (22.3) | 94 (51.1) | 43 (23.4) | 6 (3.3) | 0 (0) |
| Taking history from patients. | 186 | 57 (30.6) | 108 (58.1) | 18 (9.7) | 3 (1.6) | 0 (0) |
| Experience while performing physical examinations | 182 | 25 (13.7) | 110 (60.4) | 41 (22.5) | 6 (3.3) | 0 (0) |
| Ward procedures such as cannulation | 185 | 62 (33.5) | 86 (46.5) | 27 (14.6) | 7 (3.8) | 3 (1.6) |
| Opportunities for outpatient training | 185 | 9 (4.9) | 48 (25.9) | 52 (28.1) | 44 (23.8) | 32 (17.3) |
| Opportunities for inpatient training | 180 | 75 (41.7) | 71 (39.4) | 26 (14.4) | 5 (2.8) | 3 (1.7) |
| Opportunities for self-learning and self-study time. | 187 | 34 (18.2) | 83 (44.4) | 46 (24.6) | 16 (8.6) | 8 (4.3) |
| Opportunities for learning in the emergency department | 183 | 7 (3.8) | 53 (29) | 63 (34.4) | 47 (25.7) | 13 (7.1) |
| Opportunities for training in the laboratory | 183 | 33 (18) | 74 (40.4) | 53 (29) | 15 (8.2) | 8 (4.4) |
| Opportunities for discussing with fellow students. | 185 | 54 (29.2) | 79 (42.7) | 35 (18.9) | 11 (5.9) | 6 (3.2) |
| Teaching from nurses | 180 | 8 (4.4) | 56 (31.1) | 52 (28.9) | 26 (14.4) | 38 (21.1) |
| Teaching from Senior House officers (SHOs) | 186 | 74 (39.8) | 70 (37.6) | 19 (10.2) | 13 (7) | 10 (5.4) |
| Teaching during mini rounds. | 186 | 42 (22.6) | 88 (47.3) | 37 (19.9) | 9 (4.8) | 10 (5.4) |
| Teaching during grand rounds. | 184 | 34 (18.5) | 90 (48.9) | 40 (21.7) | 11 (6) | 9 (4.9) |
| Overall learning experience during the rotation | 187 | 20 (10.7) | 128 (68.4) | 34 (18.2) | 5 (2.7) | 0 (0) |

**Table 3.**
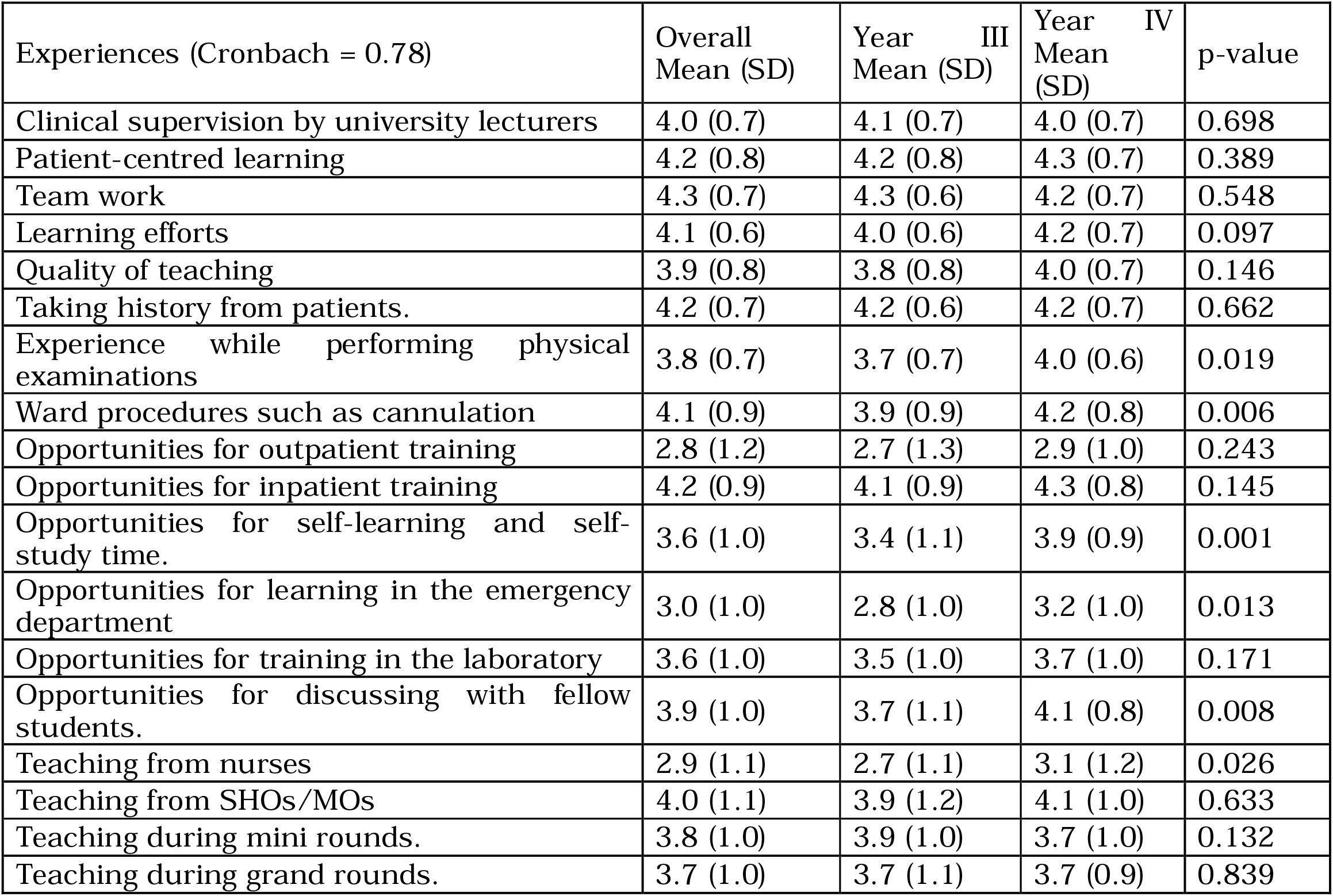
Mean experience scores of medical students during internal medicine rotation amidst COVID-19 pandemic.

| Experiences (Cronbach = 0.78) | Overall Mean (SD) | Year III Mean (SD) | Year IV Mean (SD) | p-value |
| --- | --- | --- | --- | --- |
| Clinical supervision by university lecturers | 4.0 (0.7) | 4.1 (0.7) | 4.0 (0.7) | 0.698 |
| Patient-centred learning | 4.2 (0.8) | 4.2 (0.8) | 4.3 (0.7) | 0.389 |
| Team work | 4.3 (0.7) | 4.3 (0.6) | 4.2 (0.7) | 0.548 |
| Learning efforts | 4.1 (0.6) | 4.0 (0.6) | 4.2 (0.7) | 0.097 |
| Quality of teaching | 3.9 (0.8) | 3.8 (0.8) | 4.0 (0.7) | 0.146 |
| Taking history from patients. | 4.2 (0.7) | 4.2 (0.6) | 4.2 (0.7) | 0.662 |
| Experience while performing physical examinations | 3.8 (0.7) | 3.7 (0.7) | 4.0 (0.6) | 0.019 |
| Ward procedures such as cannulation | 4.1 (0.9) | 3.9 (0.9) | 4.2 (0.8) | 0.006 |
| Opportunities for outpatient training | 2.8 (1.2) | 2.7 (1.3) | 2.9 (1.0) | 0.243 |
| Opportunities for inpatient training | 4.2 (0.9) | 4.1 (0.9) | 4.3 (0.8) | 0.145 |
| Opportunities for self-learning and self-study time. | 3.6 (1.0) | 3.4 (1.1) | 3.9 (0.9) | 0.001 |
| Opportunities for learning in the emergency department | 3.0 (1.0) | 2.8 (1.0) | 3.2 (1.0) | 0.013 |
| Opportunities for training in the laboratory | 3.6 (1.0) | 3.5 (1.0) | 3.7 (1.0) | 0.171 |
| Opportunities for discussing with fellow students. | 3.9 (1.0) | 3.7 (1.1) | 4.1 (0.8) | 0.008 |
| Teaching from nurses | 2.9 (1.1) | 2.7 (1.1) | 3.1 (1.2) | 0.026 |
| Teaching from SHOs/MOs | 4.0 (1.1) | 3.9 (1.2) | 4.1 (1.0) | 0.633 |
| Teaching during mini rounds. | 3.8 (1.0) | 3.9 (1.0) | 3.7 (1.0) | 0.132 |
| Teaching during grand rounds. | 3.7 (1.0) | 3.7 (1.1) | 3.7 (0.9) | 0.839 |

Of concern, experiences on opportunities for outpatient contacts (mean: 2.8±1.2), emergency care contacts (mean: 3.0±1.0) and teaching from nurses (mean: 2.9±1.1) were generally poor. On bivariate analysis (**Table 3**), senior clerks had better experiences in most of the items assessed. Experiences were significantly better among senior clerks than junior clerks for performing physical examinations (P= 0.019), ward procedures (P= 0.006), opportunities for self-study (P= 0.001), opportunities for peer discussions (P= 0.008), emergency care contacts (P= 0.013), and teaching from nurses (P= 0.026).

### Attitudes towards internal medicine rotation during COVID-19 pandemic

The majority of participants reported feeling safe with and having opportunity to learn from their senior or junior counterparts (Table 4). Overall, 81.7% (n=152) of the medical students felt at risk of contracting COVID-19 during the rotation and more than two-thirds (67.9%, n=127) agreed that it was difficult to follow the standard operating procedures for control of COVID-19 while clerking patients. In addition, about one-third (34.2%) felt that they did not have adequate personal protective equipment during the rotation. However, 124 participants (67.8%) agreed that they had a sufficient number of patients for them to learn from during the rotation. Notably, 86 participants (46%) agreed that assessments by tutors were difficult when senior and junior clerks were combined together. About one fifth of the medical students (11.9%, n=22) felt discouraged from pursuing a career in Internal Medicine during the rotation.

**Table 4.** Attitudes of medical students towards internal medicine rotation amidst the COVID-19 pandemic.

| Attitudes during COVID-19 (Cronbach = 0.71) | n | Strongly agree, n (%) | Agree n (%) | Neutral n (%) | Disagree n (%) | Strongly disagree, n (%) |
| --- | --- | --- | --- | --- | --- | --- |
| I have the opportunity to learn from my senior or junior counterparts. | 187 | 83 (44.4) | 86 (46) | 10 (5.3) | 6 (3.2) | 2 (1.1) |
| I feel safe around my senior or junior counterparts. | 187 | 81 (43.3) | 84 (44.9) | 15 (8) | 6 (3.2) | 1 (0.5) |
| I can comfortably share patients with my senior or junior counterparts. | 185 | 90 (48.6) | 70 (37.8) | 15 (8.1) | 8 (4.3) | 2 (1.1) |
| I felt at risk of contracting COVID-19 during the rotation. | 186 | 75 (40.3) | 77 (41.4) | 16 (8.6) | 13 (7) | 5 (2.7) |
| The patients were cooperative during my clerkship. | 186 | 56 (30.1) | 94 (50.5) | 30 (16.1) | 4 (2.2) | 2 (1.1) |
| I was able to attend all bedside teachings organized in my ward. | 185 | 76 (41.1) | 67 (36.2) | 15 (8.1) | 22 (11.9) | 5 (2.7) |
| I was able to interact with the seniors effectively during my rotation. | 186 | 42 (22.6) | 87 (46.8) | 32 (17.2) | 17 (9.1) | 8 (4.3) |
| I was able to attend online sessions organized by the lecturers. | 182 | 52 (28.6) | 73 (40.1) | 21 (11.5) | 26 (14.3) | 10 (5.5) |
| It was difficult to observe COVID-19 SOPs while clerking patients appropriately. | 187 | 46 (24.6) | 81 (43.3) | 28 (15) | 25 (13.4) | 7 (3.7) |
| Patients are always enough for all the students to clerk. | 183 | 60 (32.8) | 64 (35) | 25 (13.7) | 28 (15.3) | 6 (3.3) |
| I was able to complete all my study objectives during the rotation. | 184 | 16 (8.7) | 79 (42.9) | 41 (22.3) | 44 (23.9) | 4 (2.2) |
| The time allocated for the rotation was adequate. | 186 | 33 (17.7) | 61 (32.8) | 34 (18.3) | 46 (24.7) | 12 (6.5) |
| Assessment is difficult when senior and junior clerks are combined together. | 187 | 40 (21.4) | 46 (24.6) | 40 (21.4) | 42 (22.5) | 19 (10.2) |
| I had adequate personal protective equipment during the rotation | 187 | 19 (10.2) | 60 (32.1) | 44 (23.5) | 51 (27.3) | 13 (7) |
| I found it easy to observe COVID-19 SOPs during clinical sessions. | 180 | 15 (8.3) | 50 (27.8) | 49 (27.2) | 51 (28.3) | 15 (8.3) |
| Learning is difficult when senior and junior clerks are combined together. | 187 | 29 (15.5) | 31 (16.6) | 29 (15.5) | 64 (34.2) | 34 (18.2) |
| Teachers attend more to the junior clerks than the senior clerks. | 184 | 9 (4.9) | 34 (18.5) | 45 (24.5) | 66 (35.9) | 30 (16.3) |
| I was discouraged from pursuing a career in Internal Medicine during my rotation. | 185 | 9 (4.9) | 13 (7) | 27 (14.6) | 59 (31.9) | 77 (41.6) |

Table 5 summarizes the mean attitude scores for each of the items assessed. Overall, attitudes were moderately positive for most of the items. On bivariate analysis (Table 5), senior clerks significantly believed that the time allocated for the rotation was adequate (P<.0001) and they were able to complete their study objectives (P<.001), compared to their junior counterparts. Senior clerks also significantly believed that learning is difficult when combined with junior clerks (mean attitude score: 3.0 vs 2.5, P=0.013).

**Table 5.** Mean attitude scores for attitudes of medical students towards internal medicine rotation amidst COVID-19 pandemic.

| Experiences | N | Overall Mean (SD) | Third Year Mean (SD) | Fifth Year Mean (SD) | Man-Whitney p-value |
| --- | --- | --- | --- | --- | --- |
| The time allocated for the rotation was adequate. | 186 | 3.3 (1.2) | 2.7 (1.1) | 4.0 (1.0) | <0.0001 |
| I had adequate personal protective equipment during the rotation | 187 | 3.1 (1.1) | 3.0 (1.1) | 3.3 (1.1) | 0.102 |
| I was able to complete all my study objectives during the rotation. | 184 | 3.3 (1.0) | 3.1 (1.0) | 3.6 (1.0) | <0.001 |
| I found it easy to observe COVID-19 SOPs during clinical sessions. | 180 | 3.0 (1.1) | 2.9 (1.1) | 3.1 (1.2) | 0.344 |
| I was able to attend all bedside teachings organized in my ward. | 185 | 4.0 (1.1) | 3.9 (1.2) | 4.2 (1.0) | 0.185 |
| I was able to attend online sessions organized by the lecturers. | 182 | 3.7 (1.2) | 3.6 (1.2) | 3.9 (1.1) | 0.072 |
| I was able to interact with the seniors effectively during my rotation. | 186 | 3.7 (1.0) | 3.7 (1.1) | 3.8 (1.0) | 0.570 |
| The patients were cooperative during my clerkship. | 186 | 4.1 (0.8) | 4.1 (0.7) | 4.0 (0.9) | 0.812 |
| It was difficult to observe COVID-19 SOPs while clerking patients appropriately (R). | 187 | 3.7 (1.1) | 3.6 (1.2) | 3.8 (1.0) | 0.192 |
| I felt at risk of contracting COVID-19 during the rotation (R). | 186 | 4.1 (1.0) | 4.0 (1.0) | 4.2 (0.9) | 0.302 |
| I was discouraged from pursuing a career in Internal Medicine during my rotation (R). | 185 | 2.0 (1.1) | 1.9 (0.9) | 2.2 (1.3) | 0.445 |
| I have the opportunity to learn from my senior or junior counterparts. | 187 | 4.3 (0.8) | 4.4 (0.8) | 4.2 (0.8) | 0.140 |
| I feel safe around my senior or junior counterparts. | 187 | 4.3 (0.8) | 4.3 (0.8) | 4.3 (0.8) | 0.882 |
| I can comfortably share patients with my senior or junior counterparts. | 185 | 4.3 (0.9) | 4.3 (0.9) | 4.3 (0.8) | 0.800 |
| Teachers attend more to the junior clerks than the senior clerks (R). | 184 | 2.6 (1.1) | 2.6 (1.1) | 2.6 (1.1) | 0.638 |
| Patients are always enough for all the students to clerk. | 183 | 3.8 (1.2) | 3.8 (1.2) | 3.7 (1.1) | 0.533 |
| Learning is difficult when senior and junior clerks are combined together (R). | 187 | 2.8 (1.3) | 2.5 (1.3) | 3.0 (1.4) | 0.013 |
| Assessment is difficult when senior and junior clerks are combined together (R). | 187 | 3.2 (1.3) | 3.2 (1.3) | 3.3 (1.3) | 0.716 |

*Note: Mean attitude scores range from 1 to 5 with 1 indicating strongly disagree, hence negative attitudes and 5 indicating strongly agree, hence positive attitudes. For reversed statements (R), the higher mean attitude scores, the more negative the attitudes and vice versa.*

### Learning preferences

More than half (51%) of the medical students preferred the status quo, as it was pre-COVID-19, that is, ward based teaching and traditional face to face lectures (**Figure 1**). Whereas up to 45% preferred blended online and conventional classroom teaching, only 2% selected simulations and mannequins. Preferences did not differ significantly among junior and senior clerks (P=0.653).

**Figure 1.**
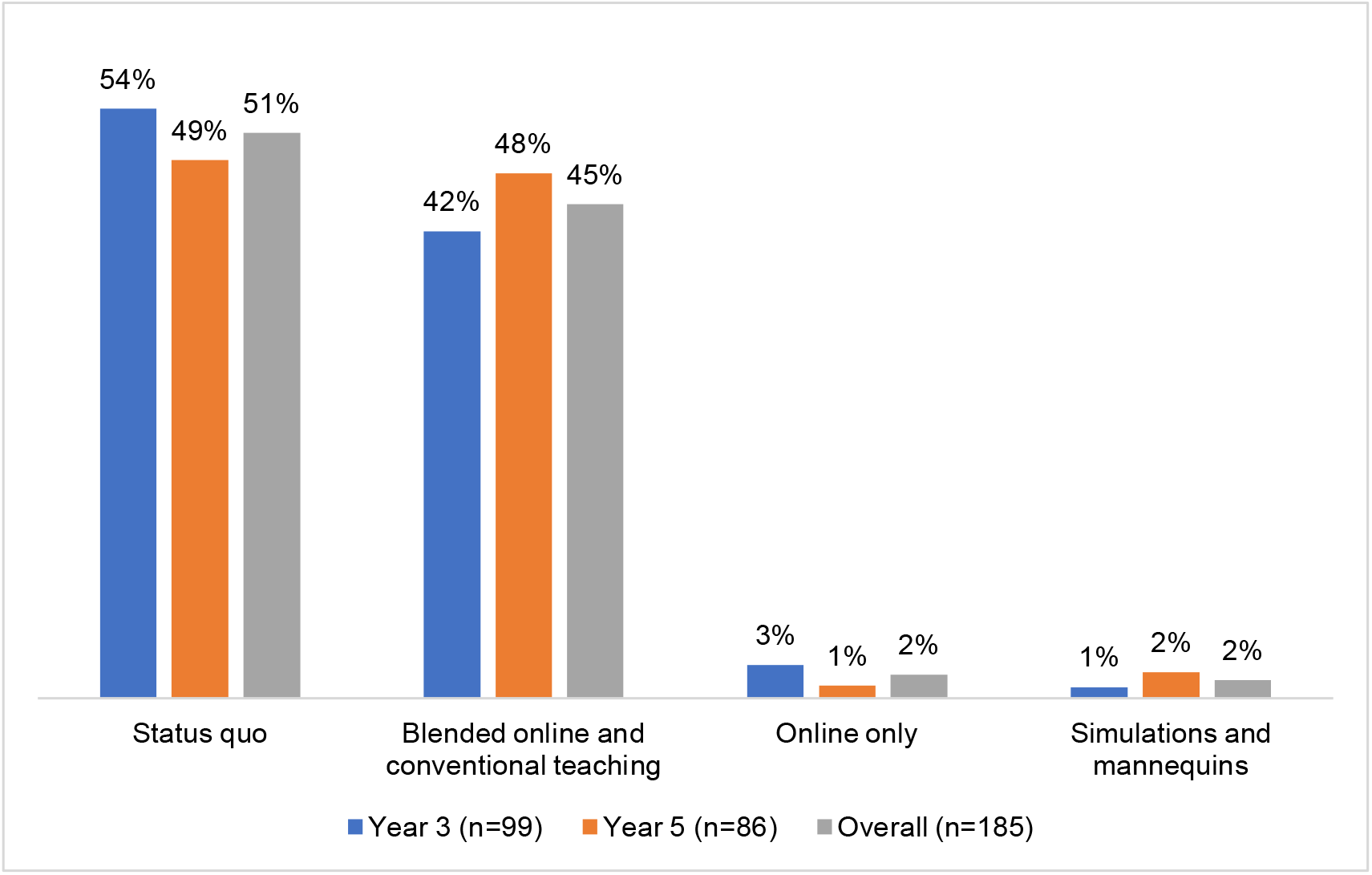
Preferences of teaching modes during the COVID-19 pandemic.

### Overall learning experience during internal medicine rotation

For the senior clerks who rotated in internal medicine in the pre-COVID era, the overall experience scores were higher before the COVID-19 pandemic compared to the rotation during the pandemic (4 vs. 3.5, P<0.0001), **Figure 2**.

**Figure 2.**
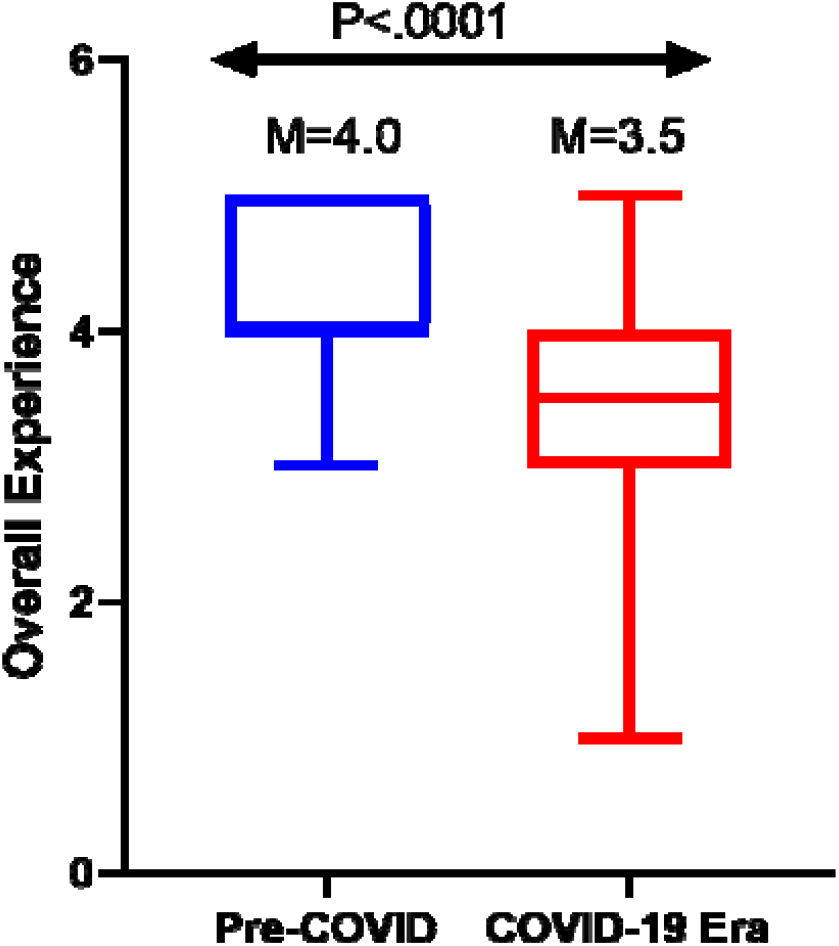
Overall experiences of medical students rotating in the Department of Medicine, Makerere University.

## DISCUSSION

Besides interrupting with clinical clerkships, the COVID-19 pandemic has brought a myriad of challenges in clinical bedside teaching since medical students now have limited direct patient care activities, such as physical examinations and procedures [11,12]. In this study, we found that nearly half of the medical students felt at risk of contracting SARS-COV-2, had difficulties in observing COVID-19 preventive SOPS, and had inadequate exposures to outpatient and emergency departments. This was consistent with anecdotal observations by the clinical supervisors during the course of the clinical rotation.

Previous studies have evaluated the impact of the COVID-19 pandemic on surgical training [13] and fellowships [14], radiology residency [15], gastroenterology residency [16] and assessment of medical students [17]. Between 40 and 100% of surgical residents reported severe impact of the COVID-19 pandemic on their training [13]. However, there is no published study examining experience of medical students or residence in general internal medicine. For the senior clerks, majority reported a good or excellent clinical experience pre-COVID-19 pandemic compared to clerkship during the pandemic.

To minimise risks of contracting SARS-CoV-2 by the learners and their teachers, many schools and universities have opted for blended online and ward teachings. Due to the negative impact of COVID-19 pandemic on medical education, there has been a shift towards e-learning at all levels of education, which may represent an optimal solution to maintain learning during this unprecedented time [18]. However, e-learning may not facilitate acquisition of clinical skills and competencies. In resource limited settings were online learning platforms are not well established it is essential that the experience of medical students is described to guide future trainings in the same setting. Despite the challenges, over half of the participants reported clinical teaching should remain as it was in the pre COVID-19 era, that is, uninterrupted clinical rotations along with class lectures. It was also interesting to note that less than 4% of the medical students opted for online only or use of simulations and mannequins as alternatives to traditional clinical teaching. This is consistent with a previous study that showed that e-learning in the clinical years is associated with low students’ satisfaction and technical challenges [18]. In Uganda, we have recently reported that medical student think e-learning is inefficient and associated with poor acquisition of knowledge [19]. Moreover, cost of data and internet stability remains a big barrier to e-learning among medical students in Uganda [19]. The use of simulations and mannequins have not been frequently implemented at Makerere University hence the low number of students who opted for it.

Notably, the costs incurred by the students during the rotation in the COVID-19 pandemic era could have had an impact on the learning experience. The government reduced the number of travellers that each public transport means was supposed to carry to enforce social distancing as a guideline for prevention of COVID-19 transmission. Costs of using public transport therefore increased to more than twice the previous costs. The means of transport provided by the university was also inadequate to cater for all the students.

The impact of interruption of clinical clerkship in internal medicine due to the COVID-19 pandemic and whether the experience of clinical clerkship during the pandemic has affected performance in theory and clinical exams of our students is not known. However, in Columbia [17], students who sat their exams during the pandemic had higher scores compared with historic cohort. However, these high scores were attributed to the open book nature of the exams and that students had more time to prepare for them.

### Study limitations

Our study had some limitations. Firstly, there was no previously standardized and validated tool to assess clinical learning experience during this COVID-19. However, we adapted an existing validated and published tool used in a similar setting but in a non-pandemic situation. Secondly, there’s paucity of published literature in this area and therefore not sufficient published studies to compare and contrast our findings with. However, to the best of our knowledge this is the first study to explore clinical learning experience of medical students in their clinical years during this pandemic in Uganda and Africa at large. Despite the above limitations, our findings provide valuable information about teaching and learning clinical medicine during the pandemic in resource-limited settings.

### Conclusions

In this study, we have reported a considerable negative effect of the COVID-19 pandemic among undergraduate medical students clerking internal medicine in Uganda. Direct patient care activities were adversely affected. Clinical teaching of junior clerks should be separate from senior clerks to maximise their clinical learning experience.

## Data Availability

The datasets analyzed during the current study are available from the corresponding author on reasonable request.

## Acknowledgment

We acknowledge Gladys Kyolaba for administrative support.

## Authors’ contribution

**Conceptualization:** Pauline Byakika-Kibwika, Felix Bongomin, Lydia Nakiyingi, Ponsiano Ocama,

**Data curation:** Ronald Olum and Felix Bongomin

**Formal analysis:** Ronald Olum and Felix Bongomin

**Funding acquisition:** Not applicable

**Investigation:** Felix Bongomin, Ronald Olum, Lydia Nakiyingi, Lalitha Rejani, Isaac Ssinabulya, Christine Sekaggya-Wiltshire, Ponsiano Ocama, Pauline Byakika-Kibwika

**Methodology:** Felix Bongomin, Ronald Olum, Lydia Nakiyingi, Ponsiano Ocama, Pauline Byakika-Kibwika

**Project administration:** Ponsiano Ocama, Pauline Byakika-Kibwika

**Resources:** Ponsiano Ocama, Pauline Byakika-Kibwika

**Software:** Felix Bongomin, Ronald Olum

**Supervision:** Lydia Nakiyingi, Ponsiano Ocama, Pauline Byakika-Kibwika

**Validation:** Felix Bongomin, Ronald Olum, Lydia Nakiyingi, Lalitha Rejani, Isaac Ssinabulya, Christine Sekaggya-Wiltshire, Ponsiano Ocama, Pauline Byakika-Kibwika

**Visualization:** Felix Bongomin, Ronald Olum, Lydia Nakiyingi, Lalitha Rejani, Isaac Ssinabulya, Christine Sekaggya-Wiltshire, Ponsiano Ocama, Pauline Byakika-Kibwika

**Writing – original draft:** Bongomin, Ronald Olum, Lydia Nakiyingi, Ponsiano Ocama, Pauline Byakika-Kibwika

**Writing – review & editing:** Felix Bongomin, Ronald Olum, Lydia Nakiyingi, Lalitha Rejani, Isaac Ssinabulya, Christine Sekaggya-Wiltshire, Ponsiano Ocama, Pauline Byakika-Kibwika

## Guarantor

Pauline Byakika-Kibwika

## Data Availability Statement

All relevant data are within the manuscript and its Supporting Information files.

## Ethical considerations

The study protocol was approved by the MakCHS School of Medicine Research and Ethics Committee (reference number #REC REF 2020-221). All participants provided written informed consent.

